# Sex differences in 277,168 arterial vascular surgical procedures: an 18-year comprehensive analysis of the Brazilian public health system

**DOI:** 10.1101/2025.09.01.25334596

**Authors:** Clara Sanches Bueno, Bruna Castelucci Bongiovanni, Nickolas Stabellini, Júlia Freire, Bruno Jeronimo Ponte, Felipe Soares Oliveira Portela, Marcelo Fiorelli Alexandrino da Silva, Marcelo Passos Teivelis, Nelson Wolosker

## Abstract

**Background:** Sex-based disparities in the presentation and outcomes of arterial vascular procedures are well-documented in high-income countries. However, comprehensive analyses within low- and middle-income healthcare systems, such as Brazil’s, are lacking. This study aimed to investigate these disparities using nationwide data from the Brazilian public health system.

**Methods:** A retrospective, nationwide analysis was conducted using data from the DATASUS repository from 2007 to 2024. It included 277,168 hospitalizations for 22 arterial vascular surgical procedures. Outcomes assessed included patient demographics, procedure frequency, age at intervention, in-hospital mortality, length of stay, and the need for intercity or interstate travel for care.

**Results:** Among 277,168 procedures, men underwent 60% of all interventions. Women were significantly older at the time of peripheral arterial interventions, while men were older for aortic and carotid procedures. Women had significantly higher mortality rates in 8 out of 22 procedures, particularly in endovascular interventions (e.g., aortoiliac stenting and extremity angioplasty. Men demonstrated a greater need to travel for surgical care, with significant differences in 14 procedures for intercity travel, and only one procedure for interstate travel (difference of approximately 1%). Regional analysis revealed the Southeast as the primary hub for specialized care, receiving patients from all other regions.

**Conclusion:** This 18-year comprehensive analysis reveals sex-based disparities in arterial vascular care within the Brazilian public health system. Men predominated in procedure volume and required more travel for care. Women presented at an older age for peripheral interventions and experienced higher mortality in specific procedures.

## Introduction

Cardiovascular diseases (CVDs), which include acute coronary syndrome, strokes, and peripheral arterial disorders (PAD), are the leading global cause of death, accounting for roughly one-third of global mortality. (1) Notably, 80 % of these fatalities occur in low- and middle-income countries. (2) In Brazil, with approximately 203 million inhabitants, CVDs have remained the primary cause of death since 1960, currently responsible for about 28.6 % of all fatalities. (3,4) PAD, encompassing peripheral atherosclerotic and carotid diseases, affects over 250 million adults worldwide, disproportionately impacting individuals aged 40 to 65 and imposing substantial morbidity as well as escalating healthcare demands, evidenced by rising numbers of vascular procedures globally. (5)

Recent global data reveal significant sex-based differences in CVD manifestations. In the United States (US), male CVD mortality rates exceed those of females. (6) Emerging evidence suggests that these sex differences also extend to therapeutic interventions, influencing outcomes in vascular surgeries. (7,8) For example, women have a higher 30-day mortality rate after lower limb revascularization. (8) They also face a greater adjusted risk of in-hospital mortality following abdominal aortic aneurysm (AAA) repair, according to a UK study involving 23.245 patients.(9) Additionally, women experience higher amputation rates in cases of critical PAD and increased graft failure rates after femoropopliteal bypass, according to analyses from the US. (10,11)

In Brazil, the public healthcare system, known as the “Sistema Único de Saúde” (SUS), is one of the world’s largest publicly funded health systems and provides complete coverage to 203 million citizens; however, in practice, 75% of this population uses the service exclusively, while 25% opt for private services. (12–14) This makes its administrative database (DATASUS) a unique epidemiological resource for assessing real-world trends in vascular care. (6,15,16) Analysis of SUS-derived data is particularly valuable due to its national representativeness, capturing socioeconomic and regional diversities that are often absent in studies from high-income countries. Additionally, the system conducts around 700,000 vascular procedures annually (7), providing a rich platform to identify sex-based disparities in understudied populations.

Although gender differences in arterial vascular procedures are well-documented in high-income countries, gaps persist in the analysis of middle-income health systems, such as Brazil. Furthermore, comprehensive studies involving large patient populations and various procedure modalities are still underrepresented in the existing literature.

## Objectives

<u>T</u>his study aimed to conduct a comprehensive assessment of sex-based disparities in postoperative outcomes following 277.168 arterial vascular procedures among patients admitted through the Brazilian public healthcare system over 18 years.

## Materials and Methods

### Data Source

All patient data were obtained from the SIH DATASUS data repository, a platform maintained by the Brazilian Ministry of Health that gathers information about hospitalizations funded by the public health system of the country (SUS)(7). The raw data underwent an ETL (extract, transform and load) process, which was created and described in the Fiocruz Applied Health Data Science platform (PCDaS), During this process, data from the SIH DATASUS regarding all hospital admissions across in the country were extracted from the platform and enriched with the integration of other databases. This resulted in a final dataset organized into multiple .csv files by state and year. Subsequently, a query was performed using Python to extract variables of interest from each table and generate a final dataset combining multiple sources. (17)

The initial data included all patients admitted from 2007 to October 2024 - data from the last two months of 2024 were unavailable at DATASUS - for 22 arterial vascular surgical procedures. These procedures are described in table 2.

The selection of cases was performed using procedure codes of the Unified Health System Table of Procedures, Medications, Orthoses, Prostheses, and Materials Management System (SIGTAP/SUS).

The study was approved by the Institutional Review Board (IRB). Since the data are unidentified on the platform, informed consent was not feasible and, as a result, was not requested by the local Ethics Committee. All analyses were performed in accordance with relevant guidelines and regulations regarding the Declaration of Helsinki.

This study compared between sexes the following analyses: the total number of procedures and patient demographics, procedures ranked by frequency, the proportion of procedures between men and women, differences in the age at which each procedure was performed, the proportion of deaths for each procedure, as well as the length of hospital stay per procedure. The difference in the need to travel between cities, states, and regions to undergo the procedure by sex was also assessed.

### Covariates

Demographics, admission, and procedure data were obtained for all eligible patients. The demographic characteristics included age, sex (male and female), race (White, Black, Yellow, Mixed, Indigenous, or others), region of residence within the country (North, Northeast, Midwest, Southeast, and South), as well as the municipality and state of residence.

Admission information included admission type (elective, urgency or other), the municipality and region of hospitalization, the date of admission, the date and status of discharge, the primary diagnosis ICD code (International Classification of Diseases). Procedure information was composed of procedure code, procedure modality (conventional or endovascular), and complexity (low, medium, or high). (18)

### Data processing and statistical analysis

Data processing and analysis were performed using R (version 4.5.0) with the Cursor integrated development environment (IDE), an AI-assisted coding platform. (19,20) This tool was employed to create and optimize statistical analysis scripts, facilitating efficient code development for large-scale data processing. Individual state files were consolidated into a unified national dataset. Data cleaning included age filtering (18-100 years), variable type standardization, and creation of key derived variables - particularly "travel_status" (comparing residence and hospitalization city/states/region.

### Variables and descriptive statistics

The study examined demographic (sex, age, ethnicity), clinical (admission type, complexity), and healthcare utilization variables (length of stay). We created three outcome metrics: 1) inter-state/city travel status, 2) discharge type (derived from mortality/billing codes), and 3) procedure complexity. Descriptive statistics included absolute/relative frequencies for categorical variables, as well as the median (with interquartile range) for continuous measures. Since the cost data obtained represent standardized reimbursement rates, no sex-based differences existed for the analyzed procedures. Consequently, this variable was excluded from our evaluation. To analyze the age difference between sexes, the Bonferroni correction was used to control for type 1 error in the presence of multiple comparisons.

## Results

This analysis included 277,168 arterial procedures, as detailed in **Table 1**. The majority occurred in men (60%), with both genders presenting nearly identical median ages (approximately 65 years). Ethnically, white patients had higher representation in men (54.2%), while females exhibited a greater Mixed-race prevalence (26.9% vs 24.3%). Most procedures were performed in the Southeast region, and the majority of admissions were emergencies (males: 64.0%; females: 63.8%). Median hospital stays were marginally shorter for women (2.9 days) than for men (3.1 days).

**Table 1.** Patient’s Demographic characteristics by gender.

| **Table 1. Patient's Demographic characteristics by gender** |  |  |
| --- | --- | --- |
| **Characteristics** | **Vascular procedures performed from 2007 to October 2024** |  |
|  | **Men** | **Women** |
| Cases - n (%) | 166,170 (60.0%) | 110,998 (40.0%) |
| Median age (IQR) | 65.1 (59-73) | 65.6 (59-75) |
| Race - n (%) |  |  |
| White | 89,994 (54.2%) | 54,004 (48.7%) |
| Black | 6,939 (4.2%) | 6,057 (5.5%) |
| Asian | 1,481 (0.9%) | 1,006 (0.9%) |
| Mixed | 40,338 (24.3%) | 29,842 (26.9%) |
| Indigenous | 53 (0.0%) | 33 (0.0%) |
| Unknown | 27,365 (16.5%) | 20,056 (18.1%) |
| Procedure Region - n (%) |  |  |
| North | 2,536 (1.5%) | 1,669 (1.5%) |
| Northeast | 22,041 (13.3%) | 19,749 (17.8%) |
| Midwest | 7,295 (4.4%) | 4,942 (4.5%) |
| Southeast | 74,838 (45.0%) | 49,215 (44.3%) |
| South | 59,460 (35.8%) | 35,423 (31.9%) |
| Travel to surgery* | 1,926 (1.2%) | 1,219 (1.1%) |
| Type of admission - n (%) |  |  |
| Elective | 59,799 (36.0%) | 40,164 (36.2%) |
| Emergency | 106,301 (64.0%) | 70,806 (63.8%) |
| Unknown | 70 (0.0%) | 28 (0.0%) |
| Length of stay - median in days (IQR) | 3.1 (1-5) | 2.9 (1-4) |
| * Interregional Travel |  |  |

**Table 2** presents the frequencies of vascular procedures performed for each gender. For men, the most common operation was intraluminal angioplasty of limb vessels without a stent (55.6%), followed by the same procedure with an uncovered stent (52.6%), and angioplasty of neck vessels or supra-aortic trunks with an uncovered stent (65.6%). Women had the same order, though with lower percentages: angioplasty without stent (44.4%), with uncovered stent (47.4%), and of neck/supra-aortic vessels (34.4%).

**Table 2.** Most common procedures for each gender.

| Procedure | Men |  |  | Women |  |  |
| --- | --- | --- | --- | --- | --- | --- |
|  | n | % | Rank | n | % | Rank |
| Intraluminal Angioplasty Of Limb Vessels (Without Stent) | 39985 | 55.6% | 1 | 31994 | 44.4% | 1 |
| Intraluminal Angioplasty Of Limb Vessels (With Uncovered Stent) | 24277 | 52.6% | 2 | 21845 | 47.4% | 2 |
| Intraluminal Angioplasty Of Neck Vessels Or Supra-Aortic Trunks (With Uncovered Stent) | 19882 | 65.6% | 3 | 10423 | 34.4% | 3 |
| Intraluminal Angioplasty Of Aorta, Vena Cava / Iliac Vessels (With Stent) | 9969 | 54.9% | 7 | 8200 | 45.1% | 4 |
| Carotid Bypass-Thromboendarterectomy | 13349 | 65.1% | 4 | 7161 | 34.9% | 5 |
| Distal Femoro-Popliteal Bypass / Thromboendarterectomy | 12432 | 66.9% | 5 | 6156 | 33.1% | 6 |
| Proximal Femoro-Popliteal Bypass / Thromboendarterectomy | 10041 | 63.7% | 6 | 5718 | 36.3% | 7 |
| Intraluminal Angioplasty Of Aorta, Vena Cava / Iliac Vessels (Without Stent) | 4326 | 55.0% | 11 | 3538 | 45.0% | 8 |
| Endovascular Repair Of Thoracic Aortic Aneurysm / Dissection With Straight Or Conical E | 5471 | 66.3% | 8 | 2784 | 33.7% | 9 |
| Distal Arterial Bypass / Thromboendarterectomy Of Other Arteries | 4622 | 64.7% | 10 | 2517 | 35.3% | 10 |
| Aorto-Femoral Bypass-Thromboendarterectomy | 3745 | 65.4% | 12 | 1977 | 34.6% | 11 |
| Infrarenal Abdominal Aortic Aneurysmectomy | 5343 | 76.7% | 9 | 1622 | 23.3% | 12 |
| Ilio-Femoral Bypass-Thromboendarterectomy | 2626 | 64.2% | 14 | 1466 | 35.8% | 13 |
| Crossover Femoro-Femoral Bypass | 3191 | 70.2% | 13 | 1354 | 29.8% | 14 |
| Intraluminal Angioplasty Of Neck Vessels / Supra-Aortic Trunks (Without Stent) | 1536 | 54.1% | 16 | 1301 | 45.9% | 15 |
| Aorto-Iliac Bypass-Thromboendarterectomy | 1149 | 64.0% | 17 | 646 | 36.0% | 16 |
| Endovascular Repair Of Iliac Aneurysm / Dissection With Tubular Endograft | 1746 | 73.7% | 15 | 622 | 26.3% | 17 |
| Intraluminal Angioplasty Of Limb Vessels (With Covered Stent) | 626 | 54.9% | 19 | 515 | 45.1% | 18 |
| Thoracoabdominal Aneurysmectomy | 697 | 66.8% | 18 | 347 | 33.2% | 19 |
| Aorto-Iliac Bifurcation Reconstruction With Angioplasty And Stents | 358 | 52.0% | 22 | 331 | 48.0% | 20 |
| Axillo-Bifemoral Bypass | 429 | 62.4% | 20 | 258 | 37.6% | 21 |
| Axillo-Femoral Bypass | 370 | 62.4% | 21 | 223 | 37.6% | 22 |

The analysis of vascular procedures, displayed in the forest plot (**Figure 1**), illustrates the adjusted odds ratios and confidence intervals for multiple vascular procedures, offering insight into their relative likelihoods. Male patients were predominant in all interventions. Among the procedures with the greatest disparity, the most notable are infrarenal abdominal aortic aneurysmectomy (23.3% female patients), endovascular repair of iliac arteries (26.3% female patients), and crossover femoro-femoral bypass (29.8% female patients), all of which show significantly lower proportions of female patients. Aortoiliac bifurcation reconstruction was the only intervention without a significant sex-based difference (48.0% female patients).

**Figure 1.**
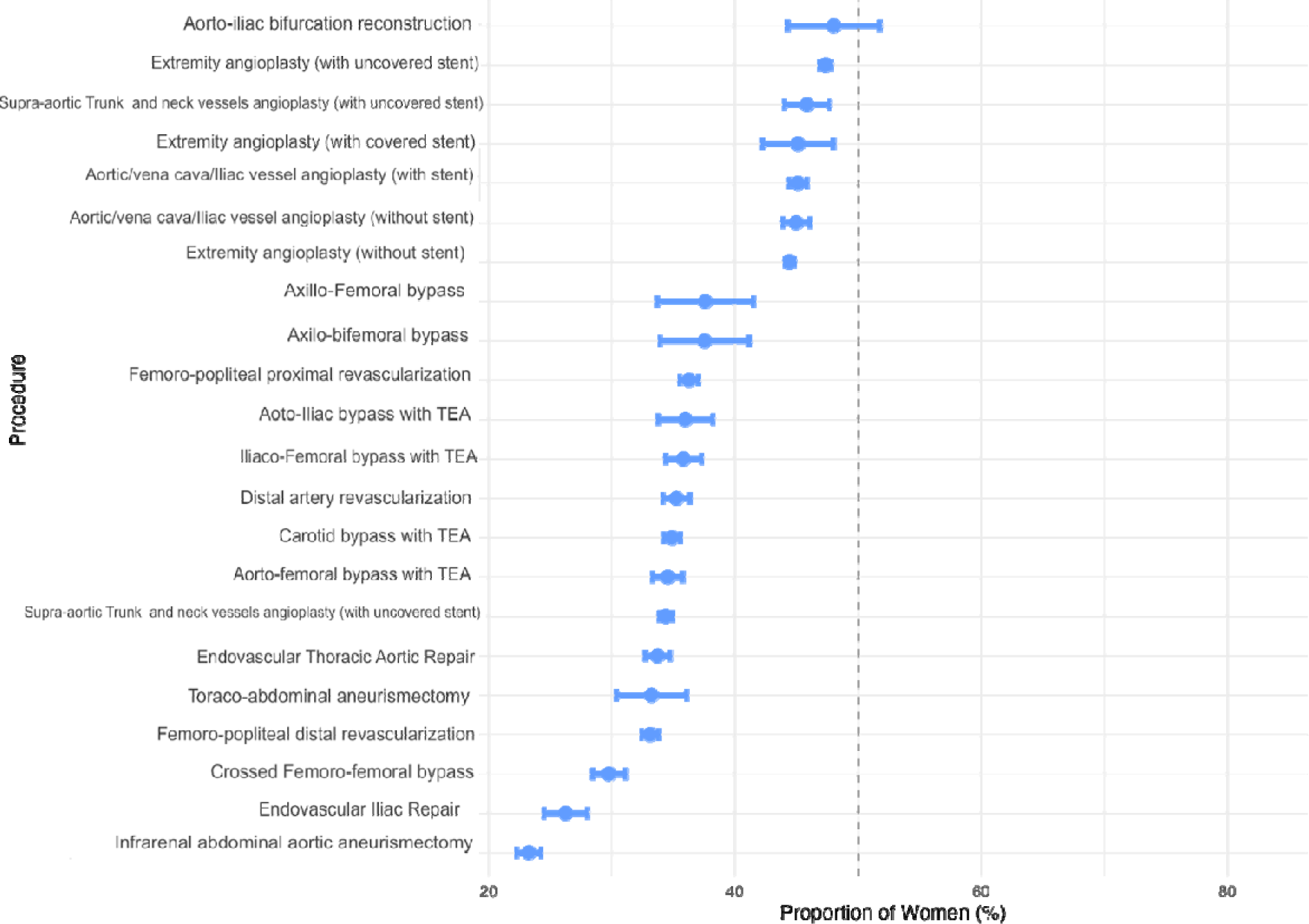
Proportion of Women by Procedure (95% CI)

**Table 3** highlights statistically significant age differences between male and female patients across various vascular procedures. Of the 19 procedures analyzed, significant differences were observed, with Bonferroni-corrected significance (***) persisting in most key comparisons, reinforcing the robustness of demographic disparities. Among the 15 procedures that retained statistical significance after Bonferroni correction, women were older at the time of intervention in 7. The largest age gap was observed in aortoiliac bypass, with men being 4.2 years older on average. These findings suggest sex-specific age trends, with women typically older when undergoing peripheral arterial interventions, while men tended to be older in aortic and carotid procedures.

**Table 3.**
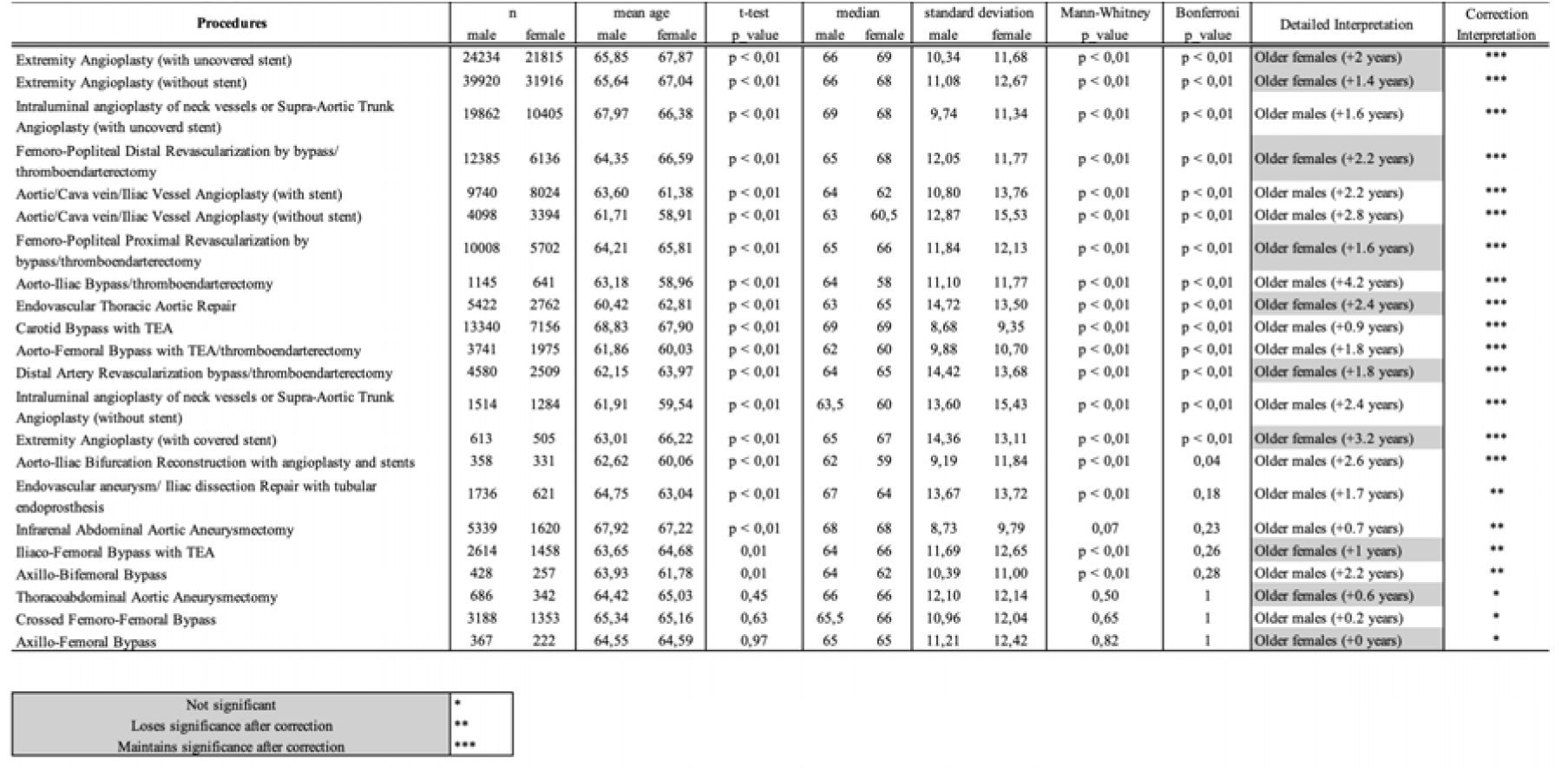
Differences on age by sex and procedure.

| Procedures | n |  | mean age |  | t-test<br>p value | median |  | standard deviation |  | Mann-Whitney<br>p value | Bonferroni<br>p value | Detailed Interpretation | Correction<br>Interpretation |
| --- | --- | --- | --- | --- | --- | --- | --- | --- | --- | --- | --- | --- | --- |
|  | male | female | male | female |  | male | female | male | female |  |  |  |  |
| Extremity Angioplasty (with uncovered stent) | 24234 | 21815 | 65,85 | 67,87 | p < 0,01 | 66 | 69 | 10,34 | 11,68 | p < 0,01 | p < 0,01 | Older females (+2 years) | *** |
| Extremity Angioplasty (without stent) | 39920 | 31916 | 65,64 | 67,04 | p < 0,01 | 66 | 68 | 11,08 | 12,67 | p < 0,01 | p < 0,01 | Older females (+1.4 years) | *** |
| Intraluminal angioplasty of neck vessels or Supra-Aortic Trunk Angioplasty (with uncovered stent) | 19862 | 10405 | 67,97 | 66,38 | p < 0,01 | 69 | 68 | 9,74 | 11,34 | p < 0,01 | p < 0,01 | Older males (+1.6 years) | *** |
| Femoro-Popliteal Distal Revascularization by bypass/thromboendarterectomy | 12385 | 6136 | 64,35 | 66,59 | p < 0,01 | 65 | 68 | 12,05 | 11,77 | p < 0,01 | p < 0,01 | Older females (+2.2 years) | *** |
| Aortic/Cava vein/Iliac Vessel Angioplasty (with stent) | 9740 | 8024 | 63,60 | 61,38 | p < 0,01 | 64 | 62 | 10,80 | 13,76 | p < 0,01 | p < 0,01 | Older males (+2.2 years) | *** |
| Aortic/Cava vein/Iliac Vessel Angioplasty (without stent) | 4098 | 3394 | 61,71 | 58,91 | p < 0,01 | 63 | 60,5 | 12,87 | 15,53 | p < 0,01 | p < 0,01 | Older males (+2.8 years) | *** |
| Femoro-Popliteal Proximal Revascularization by bypass/thromboendarterectomy | 10008 | 5702 | 64,21 | 65,81 | p < 0,01 | 65 | 66 | 11,84 | 12,13 | p < 0,01 | p < 0,01 | Older females (+1.6 years) | *** |
| Aorto-Iliac Bypass/thromboendarterectomy | 1145 | 641 | 63,18 | 58,96 | p < 0,01 | 64 | 58 | 11,10 | 11,77 | p < 0,01 | p < 0,01 | Older males (+4.2 years) | *** |
| Endovascular Thoracic Aortic Repair | 5422 | 2762 | 60,42 | 62,81 | p < 0,01 | 63 | 65 | 14,72 | 13,50 | p < 0,01 | p < 0,01 | Older females (+2.4 years) | *** |
| Carotid Bypass with TEA | 13340 | 7156 | 68,83 | 67,90 | p < 0,01 | 69 | 69 | 8,68 | 9,35 | p < 0,01 | p < 0,01 | Older males (+0.9 years) | *** |
| Aorto-Femoral Bypass with TEA/thromboendarterectomy | 3741 | 1975 | 61,86 | 60,03 | p < 0,01 | 62 | 60 | 9,88 | 10,70 | p < 0,01 | p < 0,01 | Older males (+1.8 years) | *** |
| Distal Artery Revascularization bypass/thromboendarterectomy | 4580 | 2509 | 62,15 | 63,97 | p < 0,01 | 64 | 65 | 14,42 | 13,68 | p < 0,01 | p < 0,01 | Older females (+1.8 years) | *** |
| Intraluminal angioplasty of neck vessels or Supra-Aortic Trunk Angioplasty (without stent) | 1514 | 1284 | 61,91 | 59,54 | p < 0,01 | 63,5 | 60 | 13,60 | 15,43 | p < 0,01 | p < 0,01 | Older males (+2.4 years) | *** |
| Extremity Angioplasty (with covered stent) | 613 | 505 | 63,01 | 66,22 | p < 0,01 | 65 | 67 | 14,36 | 13,11 | p < 0,01 | p < 0,01 | Older females (+3.2 years) | *** |
| Aorto-Iliac Bifurcation Reconstruction with angioplasty and stents | 358 | 331 | 62,62 | 60,06 | p < 0,01 | 62 | 59 | 9,19 | 11,84 | p < 0,01 | 0,04 | Older males (+2.6 years) | *** |
| Endovascular aneurysm/ Iliac dissection Repair with tubular endoprosthesis | 1736 | 621 | 64,75 | 63,04 | p < 0,01 | 67 | 64 | 13,67 | 13,72 | p < 0,01 | 0,18 | Older males (+1.7 years) | ** |
| Infrarenal Abdominal Aortic Aneurysmectomy | 5339 | 1620 | 67,92 | 67,22 | p < 0,01 | 68 | 68 | 8,73 | 9,79 | 0,07 | 0,23 | Older males (+0.7 years) | ** |
| Iliaco-Femoral Bypass with TEA | 2614 | 1458 | 63,65 | 64,68 | 0,01 | 64 | 66 | 11,69 | 12,65 | p < 0,01 | 0,26 | Older females (+1 years) | ** |
| Axillo-Bifemoral Bypass | 428 | 257 | 63,93 | 61,78 | 0,01 | 64 | 62 | 10,39 | 11,00 | p < 0,01 | 0,28 | Older males (+2.2 years) | ** |
| Thoracoabdominal Aortic Aneurysmectomy | 686 | 342 | 64,42 | 65,03 | 0,45 | 66 | 66 | 12,10 | 12,14 | 0,50 | 1 | Older females (+0.6 years) | * |
| Crossed Femoro-Femoral Bypass | 3188 | 1353 | 65,34 | 65,16 | 0,63 | 65,5 | 66 | 10,96 | 12,04 | 0,65 | 1 | Older males (+0.2 years) | * |
| Axillo-Femoral Bypass | 367 | 222 | 64,55 | 64,59 | 0,97 | 65 | 65 | 11,21 | 12,42 | 0,82 | 1 | Older females (+0 years) | * |

|  |  |
| --- | --- |
| Not significant | * |
| Loses significance after correction | ** |
| Maintains significance after correction | *** |

**Table 4** presents gender-based differences in mortality rates across vascular procedures. Women had significantly higher mortality rates than men in 8 out of the 22 interventions, while no significant differences between sexes were observed in the remaining 16. Notably, the highest mortality rates occurred in open aortic surgeries (thoracoabdominal: ∼41% in both sexes) and axillofemoral bypasses (11.2–17.4%), with no significant gender disparities.

**Table 4.**
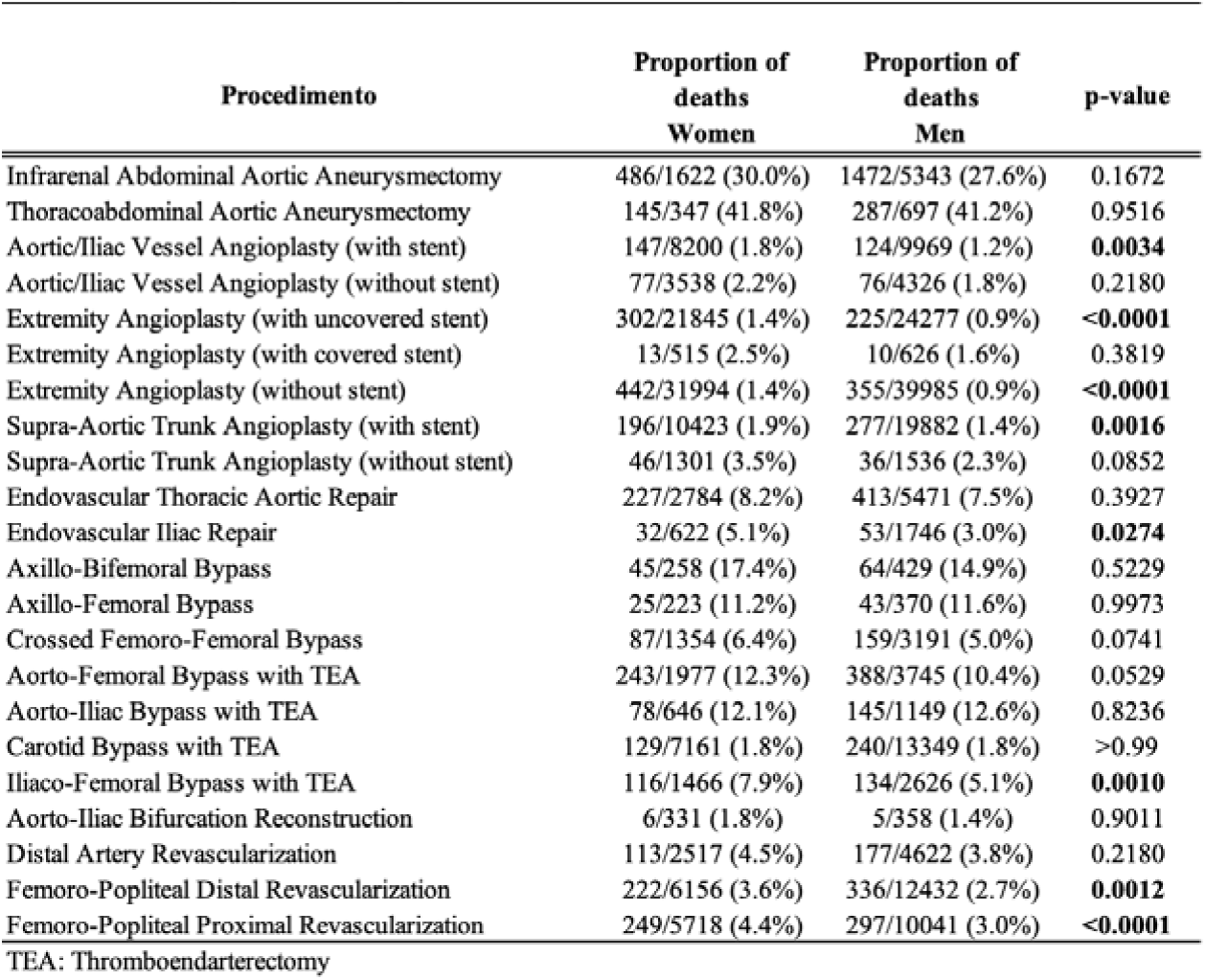
Deaths by gender for each procedure.

The analysis of hospital length of stay by gender is presented in Figure 2. For 20 vascular procedures, there were no significant differences in hospital length of stay. However, in some specific cases, women had slightly but significantly longer average hospitalization times, such as in Distal Femoropopliteal Revascularization (10.2 vs. 9.1 days). On the other hand, men experienced notably longer stays for aorto-iliac bypass with thromboendarterectomy, extremity angioplasty with a covered stent, and axillo-femoral bypass.

**Figure 2.**
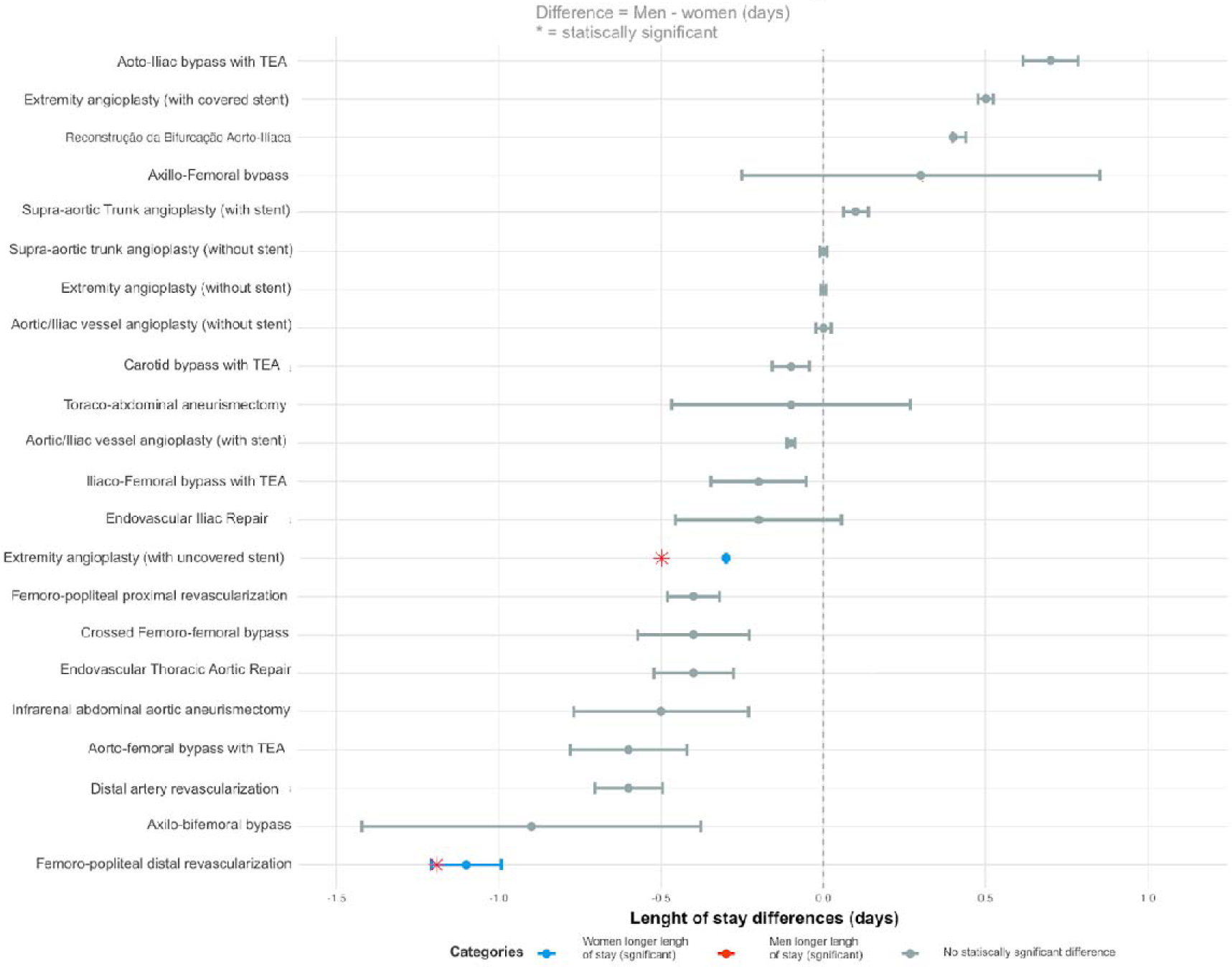
Sex differences in lenth of stay.

Table 5 and Figure 3 compare sex-based disparities in the need to travel beyond one’s municipality or state of residence for surgical procedures. Despite modest differences, men showed higher emigration rates for all vascular surgeries, with statistically significant differences in 14 of the 24 analyzed procedures for intercity travel. For interstate journeys, only one procedure showed a significant difference: iliacofemoral thromboendarterectomy (with greater male patient travel).

**Table 5.**
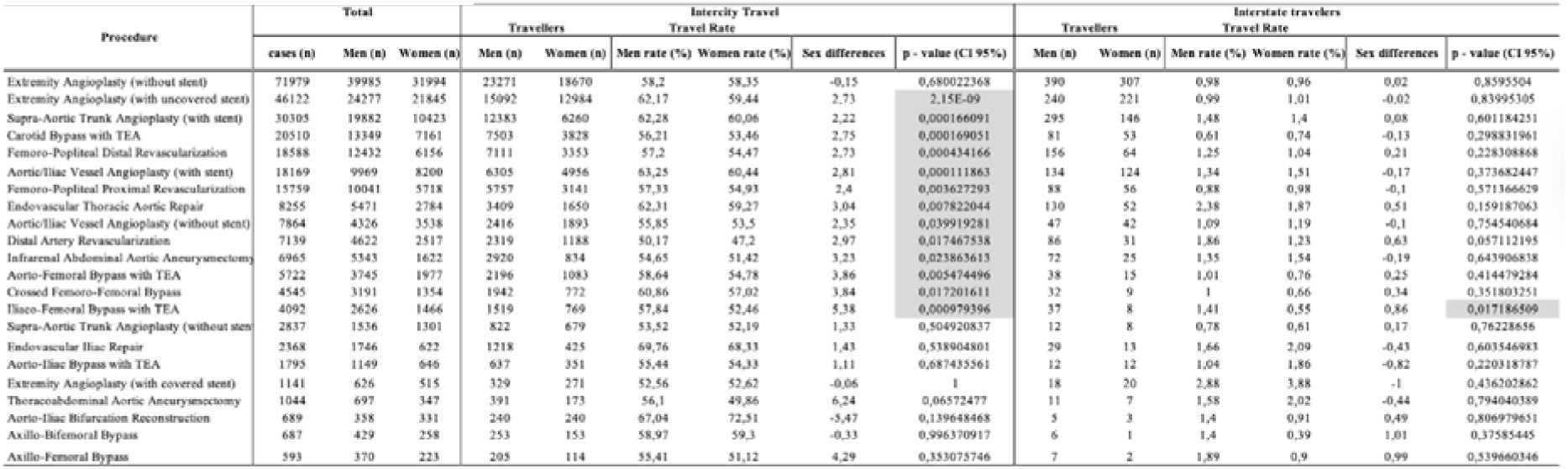
Sex disparities in intercity/interstate travel for surgery.

**Figure 3.**
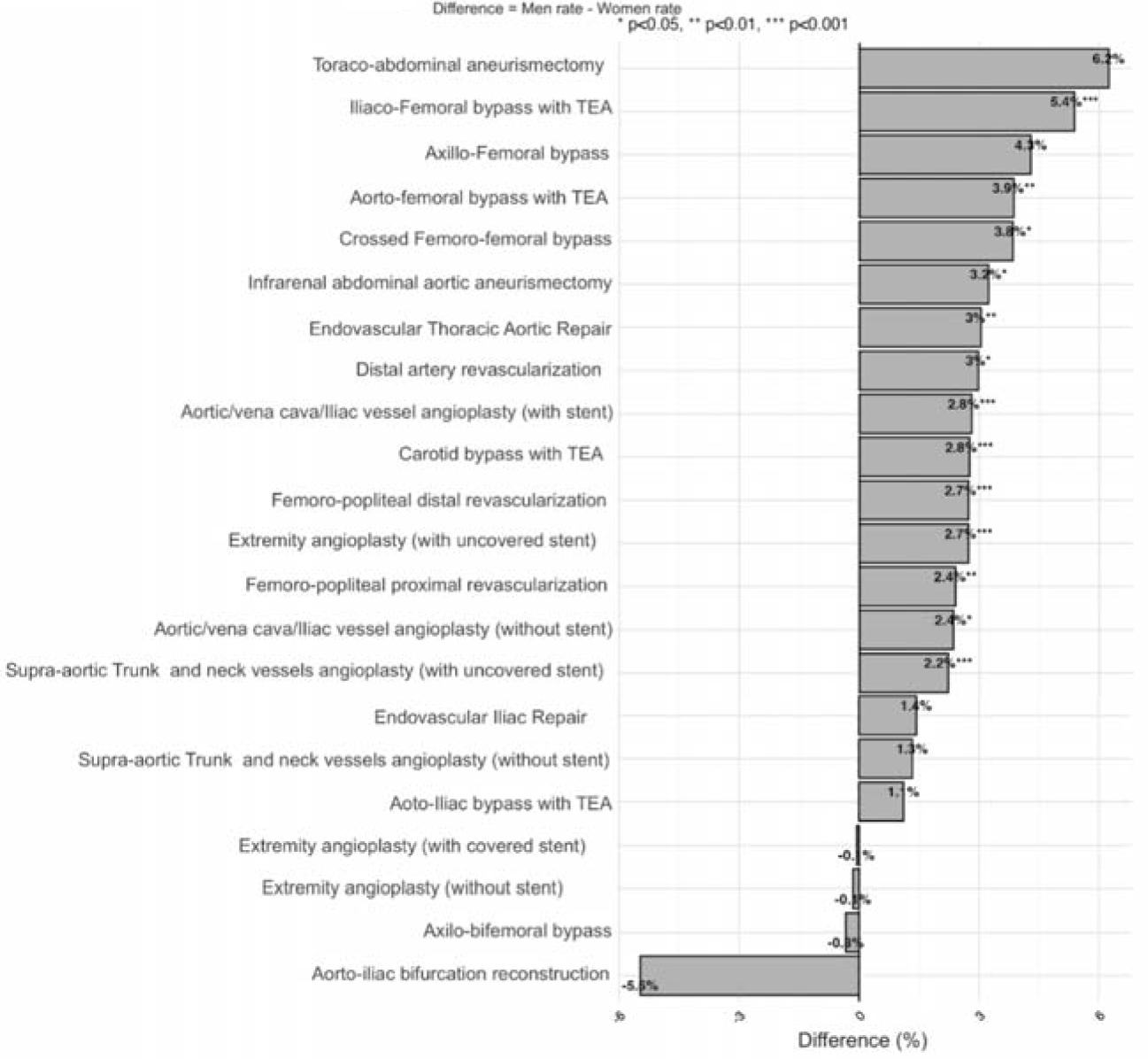
Intercity Travel differences between sexes.

The analysis of interregional surgical referrals (Table 6) revealed consistent sex-based disparities, with men demonstrating higher referral rates than women across all regions. The North region showed the highest referral rates for both sexes. The Southeast and South regions maintained uniformly low referral rates for both sexes (all <0.2%) while consistently serving as primary receiving regions for referrals from other areas.

**Table 6.** Referral region differences between sex.

| Residency region | sex | Total Referrals | Referral rate | Referral Surgery region* |
| --- | --- | --- | --- | --- |
| Midwest | Women | 50 | 1,01% | Southeast, South, North, Northeast |
| Midwest | Men | 78 | 1,07% | Southeast, South, North, Northeast |
| Northeast | Women | 100 | 0,50% | Southeast, South, Midwest, North, |
| Northeast | Men | 154 | 0,70% | Southeast, Midwest, South, North, |
| North | Women | 83 | 4,77% | South, Midwest, Southeast, Northeast |
| North | Men | 160 | 5,96% | Southeast, South, Midwest, Northeast |
| Southeast | Women | 81 | 0,16% | South, Midwest, Northeast, North |
| Southeast | Men | 126 | 0,17% | South, Midwest, Northeast, North |
| South | Women | 35 | 0,10% | Southeast, Northeast, Midwest, North |
| South | Men | 68 | 0,11% | Southeast, Midwest, Northeast, North |

\* From the most frequent referral region to the least one

## Discussion

This retrospective study utilizes a large (277,168 surgeries) population data from a publicly available dataset, TabNet (DATASUS), to assess sex differences over 18 years for 22 arterial procedures. This real-world data study provides insights into sex disparities in outcomes for vascular procedures in a continental, low- to middle-income country.

When examining demographic characteristics without procedure-specific stratification, we found that most vascular interventions were performed on white men undergoing emergency procedures in the Southeast region. This pattern reflects established sex differences in atherosclerosis progression, with men typically developing more extensive and distal arterial disease at younger ages than women. This disparity likely results from both the protective effects of estrogen in premenopausal women, which may delay atherosclerotic progression, and the higher prevalence of traditional cardiovascular risk factors in men, particularly smoking and dyslipidemia.(21–23) The concentration of procedures in the Southeast region is linked to the fact that it is the most populous region of the country, with 42% of the entire national population, and it has the highest availability of healthcare infrastructure.(24)

It is also worth noting that primary angioplasty is the most common procedure for both sexes, not only due to the prevalence of the underlying disease but also because patients may have undergone multiple procedures, since patency is not high in endovascular surgery for extremities.(25)

The observed age disparity in peripheral artery interventions - with women being significantly older than men at the time of extremity angioplasty and femoro-popliteal bypass - can be linked to well-documented biological and clinical factors. Postmenopausal estrogen depletion plays a critical role in this phenomenon. During reproductive years, estrogen’s vasoprotective effects contribute to the delayed onset of atherosclerosis and cardiovascular disease in women. However, the decline in estrogen levels during menopause results in loss of these protective mechanisms, leading to later presentation of symptomatic peripheral artery disease compared to men.(26,27)

Additionally, women’s smaller arterial anatomy may contribute to more diffuse disease patterns and greater technical challenges during revascularization, further delaying eligibility for intervention. (28–30) These biological factors are compounded by diagnostic delays, as women more frequently present with atypical symptoms rather than classic claudication, which clinicians may underrecognize, resulting in later-stage diagnosis and treatment.(31–34)

Gender disparities in cardiovascular risk and healthcare access further exacerbate this trend.(33) Studies consistently show that women undergo vascular procedures 3–7 years later than men; a gap slightly wider than the ∼2-year difference observed in our study. (35) This reflects both biological delays in disease manifestation and systemic barriers to timely care. However, the gap observed in our national cohort may be narrower because men in our setting may also experience delays in accessing the healthcare system.

Our findings in aorto-iliac disease, with men undergoing the procedure at an age ∼4-year later than woman, contrasts with existing literature from high-income countries, which typically reports younger ages for men undergoing aorto-iliac bypass. (36) This paradoxical observation, where women in our study required intervention at younger ages despite men’s higher prevalence of traditional PAD risk factors (e.g., smoking, atherosclerosis), aligns more closely with Global Burden of Disease data from low- and middle-income countries.(37) These data suggest a higher PAD burden among women, particularly younger and non-White populations, even after adjusting for conventional cardiovascular risks.(33) The sex paradox in PAD is further underscored by the disproportionate disease prevalence in women despite their historically lower smoking rates, implying sex-specific biological or socioenvironmental influences.

Cultural factors may contribute to this disparity: men in underserved regions might delay healthcare-seeking due to socioeconomic or gender norms, while women’s greater exposure to lifestyle stressors in underdeveloped countries (e.g., occupational physical demands, limited healthcare access) could accelerate disease progression. Additionally, sex differences in disease phenotype, with women more prone to diffuse arterial involvement and inflammatory pathways, may drive earlier surgical needs.(38) However, our dataset lacks risk-factor information to replicate this analysis in Brazil.

Neck vessel interventions also show a counterintuitive pattern, being more common in younger women. This trend may be associated with the higher prevalence of non-atherosclerotic diseases in this demographic, such as Takayasu’s arteritis. (39) However, as this code encompasses several distinct procedures, it is more challenging to identify a clear hypothesis for this trend.

Significant gender disparities in vascular procedure outcomes were identified, with women demonstrating higher mortality in endovascular interventions (aortoiliac stenting, extremity angioplasty, and iliac-femoral bypass) despite contrasting international data showing male-predominant or equivalent mortality. (33,40,41) This discrepancy is potentially explained by: our large real-world dataset, women’s older age at diagnosis and technical challenges (smaller vessels, distinct calcification). Paradoxically, no sex differences emerged in high-risk open surgeries (thoracoabdominal aneurysm repair, axillo-femoral bypass), suggesting procedure-specific female vulnerability rather than generalized comorbidity effects.

The finding that women had slightly longer hospitalization times for both peripheral angioplasty and distal femoropopliteal revascularization aligns with global data trends.(7,42,43) This prolonged hospitalization may occur due to women being older at the time of procedure, while also possibly accumulating more comorbidities, factors known to increase postoperative complications and delay discharge. (44) While this one-day difference may not be clinically relevant, this result remains consistent with established literature on sex differences in vascular surgery outcomes. (42,45)

Men demonstrate a greater need to travel for vascular care, with statistically significant differences observed in 14 out of the 22 analyzed procedures for intercity travel. This pattern may reflect a probable concentration of specialized surgical resources for common complex procedures within major urban centers within the same state. Given their generally higher prevalence of vascular disease, men might more frequently require these services and could consequently be referred to them more often. In contrast, interstate travel, which showed a male predominance in only one procedure, is likely reserved for highly exceptional, ultra-specialized care. (46) The overall absence of a clear gender disparity in this context suggests that the need to be referred could be driven primarily by extreme clinical complexity and the pursuit of unique expertise available at specific centers, rather than by patient gender.

Brazil’s intense regional disparities have led to patient migration for vascular surgical procedures. However, most cases involve intraregional flows within the same geographic region; interregional referrals were also documented, revealing distinct patterns of healthcare-seeking behavior. The Northern region exhibited the highest referral rates, primarily directed toward the Southeast, a technological hub with advanced medical infrastructure. This was also the case for the South and Midwest regions, likely due to geographic proximity. In contrast, the Southeast and South consistently functioned as the primary receiving regions for interregional referrals, reflecting their greater concentration of specialized vascular services. (4)

A sex-based analysis of interregional referrals revealed that men more frequently travelled to another region to seek out surgical care, and in most cases, both men and women were primarily referred to the same region, the Southeast. This suggests that there are underlying factors determining referral patterns — such as availability of specialized services or geographic accessibility — affecting both sexes similarly, concentrating their destination to the region with the most resources regarding access to vascular surgical care.(47) An exception to be noted is that women from the Northern region were mostly referred to the South and Midwest, differing from the data seen across all other regions. Further assessment is necessary to understand the reasons for this discrepancy, as there was no clear explanation found within this study.

## Limitations

This study has inherent limitations due to the use of aggregated administrative database TabNet (DATASUS), which are subject to coding errors, reporting inconsistencies, and potential misclassification or underreporting. Data anonymization prevented longitudinal tracking of individual patients, so unique patient counts cannot be distinguished from total procedures, as one patient may have undergone multiple surgeries.

Also, inconsistencies in coding and reporting quality made it impossible to categorize patients by comorbidities or clinical severity, limiting risk-factor analysis. A methodological limitation arises from procedure codes encompassing multiple interventions with substantially distinct epidemiological profiles and surgical risk levels, potentially introducing interpretation challenges and confounding effects in outcome analyses.

Nevertheless, these limitations are counterbalanced by significant strengths. This study consolidates Brazil’s public health system records, providing a comprehensive real-world analysis of sex-disparities in arterial vascular care.

## Conclusion

This nationwide retrospective study of 277,168 arterial procedures revealed significant sex-based disparities: men predominated in surgeries (66.17%), reflecting higher prevalence of arterial risk factors, while women underwent peripheral interventions at older ages and showed higher mortality in endovascular procedures. Regional inequalities show predominant patient migration from the North to Southeast/South regions, indicating concentration of specialized resources. Although men travelled more frequently between regions, referral patterns were similar between sexes.

## Data Availability

All data produced in the present study are available upon reasonable request to the authors

https://datasus.saude.gov.br/acesso-a-informacao/producao-hospitalar-sih-sus/

